# Potential use of Immunodaat® (Botanical extract of Elderberry -Sambucus Nigra L.) in the management of Post Covid-19 symptoms- a comparative, multi-centric, randomized, clinical study

**DOI:** 10.1101/2022.10.04.22280680

**Authors:** Shailesh Deshpande, Narendra Mundhe, Vaishali Deshpande, Sanjay Tamoli, Swapnali Mahadik, Vinay Pawar

## Abstract

**Background:** Immunodaat™, a proprietary botanical extract, contains standardized extract of Elderberry (*Sambucus Nigra L*.). Elderberry is used as an immunity enhancer for prevention and management of various respiratory diseases and as an anti-oxidant.

**Objectives:** The present study evaluated the effect of Immunodaat™, in Post Covid-19 recovery.

**Methods:** An open labeled, comparative, randomized, multi-centric, prospective clinical study was conducted on 74 subjects who had post COVID-19 symptoms recovery from mild-moderate COVID-19 within the last 15 days. Subjects were divided equally, with 37 subjects in trial and control groups each, where subjects in trial group were given Immunodaat™ capsules in a dose of 250 mg Capsules twice daily along with the standard care for COVID-19 for consecutive 30 days while subjects in control group were given only the standard care for Post Covid. Subjects were evaluated for post clinical recovery signs and symptoms (like cough, fatigue, Myalgia, Joint pain, confusion, altered mood, anxiety, insomnia etc.) and lab parameters. Subjects were evaluated for quality of life using WHO QOL BREF.

**Results:** Continuous 30 days administration of Immunodaat™ showed a significant difference in reducing physical as well as mental symptoms when compared to the control group. Also, a significant improvement in quality of life was observed on WHO QOL BREF scale with the use of Immunodaat compared to the control group. Overall quality of life, energy levels and stamina levels improved with the use of Immunodaat™.

Subjects showed excellent tolerability and did not have any adverse effect with the use of Immunodaat™.

**Conclusion:** The study concludes that use of Immunodaat™ over 30 days aided to normalize the physical and mental symptoms that occurred due to Post COVID and long COVID. Immunodaat™ can be considered as a safe and effective natural ingredient in the management of Post COVID-19 or long COVID condition.

## Background information (Introduction)

The COVID-19 pandemic has had devastating effects on the global population. Since the beginning of the pandemic, the scientific community has worked tirelessly to understand the diagnosis, pathogenesis, and management of this disease. However, there are limitations in our understanding of the disease, and therefore managing COVID-19 is still a major challenge. While most COVID-19 infections remain asymptomatic or mild, the possibility of the disease progressing to the severe stage, with the possibility of mortality, remains between 0.5 to 5%.^1-2^It has been observed that even those who have recovered from the disease may continue to have both physical and psychological symptoms, which may be present for weeks or months after the formal disease state has ended. The SARS-Cov-2 virus can result in damageto the lungs, heart, and brain, all of which increases the risk of long-term health complications.^3-4^

The United States Center for Disease Control (CDC) defines **post-COVID conditions** as a wide range of new, returning, or ongoing health problems that individuals may experience for **four or more weeks** after initial infectionfrom the virus that causes COVID-19. These conditions can present themselves as a variety of health complications with varying disease durations. These post-COVID conditions may also be known as long COVID, long-haul COVID, post-acute COVID-19, long-term effects of COVID, or chronic COVID. In this paper, we will refer to this post-COVID-19 condition as “long COVID.” Common signs and symptoms of long COVID include fatigue, shortness of breath/ difficulty breathing, cough, and/orchest pain. Psychological symptoms may include impaired memory, impaired concentration, depression, anxiety, and/or sleep disturbances.Other complications may includemuscle pain, joint pain, headache, fast or pounding heartbeat, loss of smell or taste, skin rashes, and dizziness.Symptoms may worsen after physical or mental activities. Some individuals may experience fever for an extended period of time. Very few people may experienceheart complications, chronic kidney impairment, stroke, and Guillain-Barre syndrome^5^Long COVID complications may occur even if the infected individual only had mild or moderate COVID symptoms. Individuals with advanced age and those with comorbid conditions are the most likely to experience lingering COVID-19 symptoms, however even young, otherwise healthy people can feel unwell for weeks to months after infection.^6^

Management of long COVID involves medical, psychological and social support to enable the Covid-19 recovered patient to return to their previous quality of life. While symptomatic management is the traditional course of treatment, other measures include: use of nutritional supplements and/or a protein rich diet. These lifestyle changes may replenish the lost nutritional status of the individual. Continued use of anti-coagulants, small doses of hypoglycemic agents, anti-depressants, and sedatives have also been recommended.7

After a COVID-19 infection, one of the majorly impacted body systems is the immune system. Herbal ingredients play a vital role in immune compromised conditions due to their high degree of safety and efficacy. The phyto-constituents in these plant materials act on different pathways to bring about enhanced ability of the body to fight infections and allow for early recovery from infections.

From the wide range of herbs being used for their immune enhancing potential, Elderberry (*Sambucus NigraL*.) has shown particular promise. In traditional and folklore medicine, elderberry has been used as an immunity enhancer for prevention and management of respiratory health, including supporting populations who had colds and influenzas. Historically, various parts of the Elderberry plant, especially its flowers and dried fruits, have been used both as foods and as remedies for health concerns. Elderberry contains anthocyanins, a subset of flavonoids, which have immuno-modulating, anti-inflammatory, and anti-oxidant effects. In this study, the elderberry fruit was utilized.

Elderberry is rich in phyto-nutrients along with minerals, vitamins and essential oils. Polyphenols, a type of phytonutrient, are present in relatively high concentrations and are known for their free radical scavenging (antioxidant) activity. In recent years, these polyphenolswere found to have antibacterial, antiviral, antidepressant activities.^8^

Immunodaat® is a proprietary branded botanical ingredient extract developed by Lodaat Pharma and composed of Elderberry extract. Based upon the antioxidant activities of Elderberry, and the composition of Immunodaat®, a hypothesis was postulated that Immunodaat®is useful in immunity support andin long COVID recovery. To test the hypothesis, a clinical study titled “A Prospective, Randomized, Open Label, Two arm, Comparative Clinical Study to evaluate the effect of Immunodaat® Botanical Ingredient, in Post Covid-19 recovery” was conducted. This report provides details of the study outcomes.

## Materials and Methods

### Study Sites, IEC approval and CTRI registration of the study

The study was conducted at two sites in India. KVTR Ayurved Institute and Hospital, Boradi, Maharashtra, India and Parul Ayurveda Institute and Hospital, Vadodra, Gujrat, India. IEC approval was obtained from both the sites and the study was registered with CTRI with no – CTRI/2021/06/034433 [Registered on: 29/06/2021]

### Study Design

The study was an open labeled, comparative, randomized, multicentric, prospective clinical study where subjects in one group were givenImmunodaat® capsules along with the standard care for COVID-19 while subjects in the other group were given only the standard care.

### Study objectives and Outcomes

Theobjectiveofthestudywas toevaluate efficacy and safety of Immunodaat®, a proprietary botanical ingredient extract from elderberry (*Sambucus nigra L*.), in long COVID Recovery.

The primary study outcome was to evaluate comparative changes in post-clinical recovery from COVID-19 (signs/symptoms/lab parameters) over a period of 30 days between the two groups. In addition, a comparative assessment on blood related parameters, including hematology, ESR and CRP from baseline to 30 days was performed between the groups.

The secondary study outcomes were to assess changes in post-clinical recovery symptoms including stress, anxiety, appetite, digestion, sleep, regularity/consistency of bowel movements, physical energy, and stamina over a period of 30 days between the two groups. Assessment of changes in health status assessment on WHO-QOL BRIEF over a period of 30 days between the two groups was performed along with Global assessment of overall change as per the study participants and the study investigator. Safety and tolerability of the study products was assessed by evaluating adverse events, both clinically as well as based on laboratory investigations.

### Sample Size

The total sample size in the study was 74 subjects. The subjects were divided equally, with 37 subjects in each of the two groups. All 74 subjects completed the study and there were no drop outs.

### Inclusion & Exclusion Criteria

#### Inclusion Criteria

Male and female subjects between the age groups of 18 and 60 years (both inclusive) who were ready to provide written informed consent, and those who had post COVID-19 symptoms after having recovered from mild to moderate COVID-19 within the last 15 days were recruited into the study. Mild to moderate symptoms of COVID-19 and that of long COVID-19 were defined as per the CDC guidelines (as outlined in www.cdc.gov/coronovirus/2019) All participating subjects were required to follow medical and health instructions provided by the regulatory authorities.

#### Exclusion Criteria

Subjects who had severe COVID 19 symptoms and severe long COVID complications were excluded from the study. Subjects who had chronic, severe, unstable, and/or uncontrolled co-existent medical illness such as hypertension, cardiovascular diseases, liver diseases, kidney diseases, lung diseases or other disease of concern were excluded. Disease of concern was defined as any disease which may place the patient at increased risk during the study. Pregnant and breast-feeding women were also excluded. Immune compromised patients, and patientshaving any type of severe allergy to herbal products were excluded from the study.

### Study Product, dosage and duration

As per acomputer-generated randomization list, subjects were randomized either to Group A i.e. Immunodaat®Capsule + Conventional management (Standard of Care) or Group B i.e.

Conventional management (Standard of Care-Vitamins like B Complex, Vitamin C, Zinc Supplements or medications as per symptom requirement.). Subjects were given the Immunodaat® Capsule in a dose of 1 Capsule (250 mg) twice daily along with their ongoing Conventional management (in Group A), subjects in the Group B were asked to continue with the on-going Conventional management for 30 days.

The key ingredient of Immunodaat® Capsules isa proprietary Elderberry Fruit Dry extract 7:1 (*Sambucus nigra L*).

### Study Procedure

The study was initiated after receiving IEC approval & subsequent registration of the study on CTRI. Subjects who had mild to moderate Covid-19 and had recovered clinically from COVID-19 were screened for eligibility criteria. Eligible subjects were screened after written consent was received. At the time of the screening visit, demographic data and medical history of the subjects were recorded. Subject’s physical & systemic examinations were performed. This evaluation included a vital exam. The subject was advised to undergo the following investigations: CBC, ESR, Hb%, CRP, Liver function tests, Renal function tests, fasting blood and post prandial blood sugar levels and urine pregnancy test (only if the subject was female of child bearing age). At baseline visit, subjects were recruited upon meeting inclusion/exclusion criteria. As per computer generated randomization list, subjects were either randomized one of the two study groups.

General and systemic examinations (including vitals) were performed. Subjects were evaluated for post clinical recovery signs, symptoms, and lab parameters. Lab parameters include cough, low grade fever, fatigue, shortness of breath, chest pain, headache, neurocognitive difficulties, muscle pains/weakness, gastrointestinal upset, skin rashes, metabolic disruption, depression and other mental health conditions. These signs and symptoms were graded as mild, moderate or severe by the investigator. Subjects were also evaluated for stress, anxiety, appetite, digestion, sleep, bowel movements, physical energy, and stamina. These were evaluated on a scale 0 to100 where 0 indicated no symptoms and 100 indicated maximum symptoms. Subjects were evaluated for quality of life using WHO QOL BREF.

In addition to the baseline visit, subjects were instructed to return to the study center on day 15 and day 30. On day 15, subjectsunderwent general and systemic examinations, including vitals. Subjects were evaluated for post clinical recovery signs, symptoms, and lab parameters. On day 30, subjects were evaluated for quality of life using WHO QOL BREF. Subject’s global evaluation for overall change and investigator’s global evaluation for overall change was performed. Tolerability of the study products was assessed by investigator and by subject. All the subjects were closely monitored for any Adverse Events from the baseline visit until the final study visit. During the final follow up visit (i.e. Day 30), subjects underwent the same laboratory investigations that were conducted at baseline. After completion of 30 days of study treatment, all the subjects were asked to stop consuming trial product. Subjects were advised by medical professionals for further treatment.

## Observations and Results

The study consort chart provides details from screening to completion in the study. The average age of subjects in the Immunodaat® group was 40.54 ±12.05 while in the control group it was 37.24± 9.57 (non significant difference, *p>*.*05*). There were 25 male subjects and 12 female subjects in Immunodaat® group while the respective numbers were 19 and 18 in the control group (non significant difference,*p>*.*05*). Vital signs including pulse, temperature, blood pressure, and respiration rate were within normal range for all subjects at the baseline visit and there was no significant difference between the two groups. Laboratory investigations including complete blood count, liver function tests, kidney function tests, lipid profile, urine analysis, blood sugar levels, ECG, and chest X-Raywere normal at baseline visits in both groups without any statistically significant difference. Table 1 Provides details.

**Table 1:** Assessment of Baseline Demographics.

| Parameter | Immunodaat® Group (n=37) | Control Group (n=37) | P value between the groups |
| --- | --- | --- | --- |
| Mean Age SD | $40.54 \pm 12.05$ | $37.24 \pm 9.57$ | |
| Gender Male | 25 | 19 |  |
| Gender Female | 12 | 18 |  |
| Pulse | $86.85 \pm 11.83$ | $84 \pm 7.14$ | |
| Temperature | $98.05 \pm 0.67$ | $98.04 \pm 0.57$ | |
| Blood Pressure (Systolic) | 118.23±7.96 | 119.18±11.15 | $p>0.05$ for all the parameters |
| Blood Pressure (Diastolic) | 75.64±6.52 | 72.21±6.17 |  |
| Respiration Rate | 19±1.01 | 20.45±2.00 |  |
| Blood Sugar Random | 102.54±23.85 | 100.14±16.67 |  |
| Hb% | 12.83 ±1.71 | 12.43±1.71 |  |
| WBC | 6986.48 ± 1367.31 | 6713.51±1545.45 |  |
| ESR | 15 ±7.23 | 11.71 ±9.55 |  |
| Total Bilirubin | 0.53 ±0.24 | 0.65 ±0.23 |  |
| SGOT | 24.83 ±7.09 | 23.08±6.73 |  |
| SGPT | 23.24±7.66 | 25.89±8.72 |  |
| Total Proteins | 7.04 ±0.82 | 7.16 ±0.70 |  |
| AG Ratio | 1.65 ±0.43 | 1.81 ±0.54 |  |
| Sr. Creatinine | 1.05 ±0.24 | 1.08 ±0.24 |  |
| BUN | 24.11 ±5.71 | 26.35±6.24 |  |
| Sr. Cholesterol | 189.31±16.48 | 187.83±23.44 |  |
| Sr. Triglycerides | 118.11±34.10 | 115.43±29.15 |  |
| HDL | 38.27±8.07 | 38.40±7.81 |  |
| LDL | 126.74±13.05 | 128.33±22.40 |  |

### Assessment of change in the Symptoms of Post COVID-19 over 30 days of study

Table 2 provides details of the symptoms of Post COVID-19 in subjects in the two groups. It was observed that there was no significant difference (*p*>.05) in the number of subjects who had symptoms of Post COVID-19 at baseline visit in the two groups. However, a significantly higher number of subjects continued to have these symptoms in the control group, compared to those takingImmunodaat® capsules. A significantly lower number of subjects in Immunodaat® group showed symptoms likefatigue, myalgia, weakness, joint pain, mental confusion, dizziness, alteration of mood, insomnia and anxiety at the end of the study. Cough, Headache, GI disturbance, Palpitation, and Shortness of Breathwere observed in a few subjects at the end of the study in both the groups and the difference was found to be non-significant.

**Table 2:** Assessment of Symptoms over 30 days Post COVID-19 recovery.

| Sr. No | Symptom | Immunodaat® Group (n=37) |  | Control Group (n=37) |  | p value between the groups |
| --- | --- | --- | --- | --- | --- | --- |
|  |  | Baseline | 30 Days | Baseline | 30 Days |  |
| 1 | Cough | 9 (24.32%) | 1 (2.70%) | 8 (21.62%) | 2 (5.40%) | >0.05 |
| 2 | Fatigue | 26 (70.27%) | 4 (10.81%) | 28 (75.67%) | 16 (43.24%) | <0.05 |
| 3 | Headache | 12 (32.43%) | 2 (5.40%) | 13 (35.13%) | 5 (13.51%) | >0.05 |
| 4 | Myalgia | 28 (75.67%) | 4 (10.80%) | 29 (78.37%) | 15 (40.54%) | <0.05 |
| 5 | Weakness | 31 (83.78%) | 5 (13.51%) | 29 (78.37%) | 14 (37.83%) | <0.05 |
| 6 | GI Disturbance | 18 (48.64%) | 5 (13.51%) | 14 (37.83%) | 7 (18.91%) | >0.05 |
| 7 | Palpitation | 19 (51.35%) | 6 (16.21%) | 19 (51.35%) | 10 (27.02%) | >0.05 |
| 8 | Joint Pain | 9 (24.32%) | 2 (5.40%) | 8 (21.62%) | 5 (13.51%) | <0.05 |
| 9 | Confusion | 12 (32.43%) | 4 (10.80%) | 14 (37.83%) | 8 (21.62%) | <0.05 |
| 10 | Dizziness | 10 (27.02%) | 1 (2.70%) | 11 (29.72%) | 5 (13.51%) | <0.05 |
| 11 | Altered mood | 12 (32.43%) | 3 (8.10%) | 13 (35.13%) | 8 (21.62%) | <0.05 |
| 12 | Insomnia | 18 (48.64%) | 3 (8.10%) | 17 (45.94%) | 8 (21.62%) | <0.05 |
| 13 | Anxiety | 14 (37.83%) | 2 (2.70%) | 13 (35.13%) | 8 (21.62%) | <0.05 |
| 14 | Dyspnea | 6 (16.21%) | 0 | 4 (10.80%) | 2 (5.40%) | >0.05 |

### Assessment of change in the Symptoms of Post COVID-19 over 30 days of study based on their severity

Post COVID-19 symptoms were also assessed on a severity scale. Measures on the scale were no symptoms, mild, moderate, and severe symptoms. It was observed that a significant reduction was observed on symptoms including fatigue, headache, myalgia, weakness and insomnia with the use of Immunodaat® as compared to control group. Symptoms including cough, GI disturbances, and shortness of breath (dyspnea) did not show any difference between the two groups. Details in Table 3.

**Table 3:**
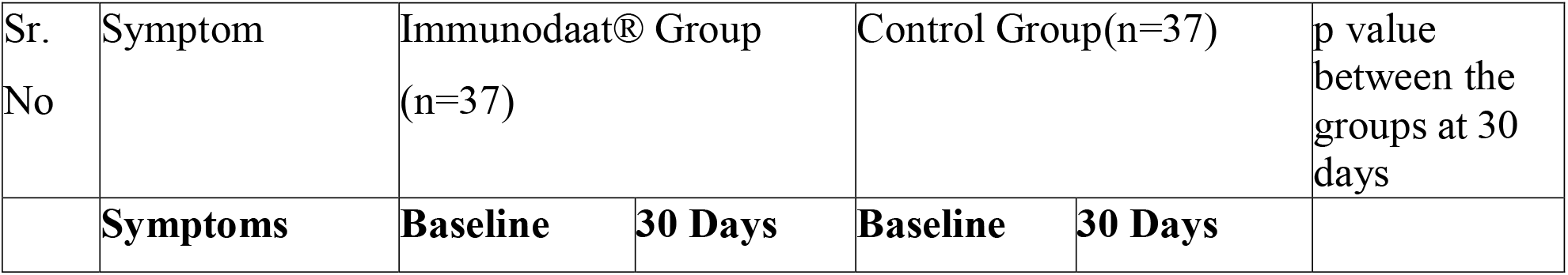
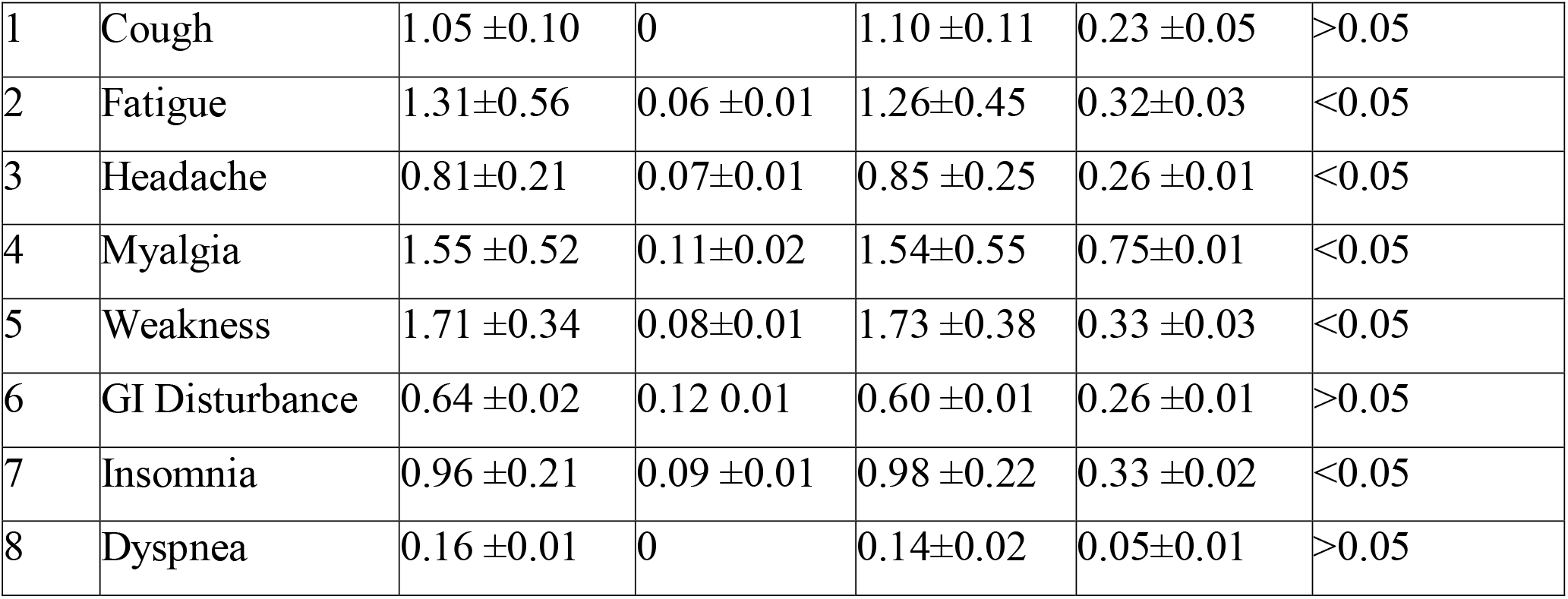
Assessment of Post Covid-19 Symptoms based on severity.

### Assessment of effect on mental health symptoms post COVID-19

Common mental health symptoms like stress, anxiety, and difficulty concentrating were assessed at baseline and at the 30 day follow-up visit. It was observed that there was a significant reduction (*p*<.05) on these parameters in both the study groups over 30 days duration. Comparative assessment between the groups observed that the reduction in Immunodaat® group was significantly greater (*p*<.05)when compared to control group. Further details can be found in Table 4.

**Table 4:** Assessment of Symptoms related to Mental Health Post Covid-19 on VAS (0-100)

| Sr. No | Symptom | Immunodaat® Group(n=31) |  | Control Group (n=30) |  | P value between groups at day 30 |
| --- | --- | --- | --- | --- | --- | --- |
|  | Parameter | Baseline | 30 Days | Baseline | 30 Days |  |
| 1 | Stress | 61.34 ±10.13 | 16.57 ±8.11* | 63.03 ±10.34 | 31.87 ±5.15* | <0.05 |
| 2 | Anxiety | 51.53 ±8.18 | 11.34 ±3.23* | 56.18 ±8.91 | 25.35 ±6.35* | <0.05 |
| 3 | Lack of concentration | 35.41 ±7.64 | 6.80±2.15* | 34.77±7.80 | 14.34±3.14* | <0.05 |

Assessment of symptoms of Stress, Anxiety and Lack of Concentration on onVAS scale ranging from of 0-100 where 0 is no symptom and 100 is maximum symptom (\*p<0.05 Significant)

Similarly, assessment of severity of Insomnia was conducted using the Insomnia Severity Index. There was a significant reduction (*p*<.05) in Insomnia in both the groups as compared to the baseline. However, the reduction in Insomnia with the use ofImmunodaat® was found to be significantly (*p*<.05)greater as compared to control. Refer to Table 5 for further details.

**Table 5:**
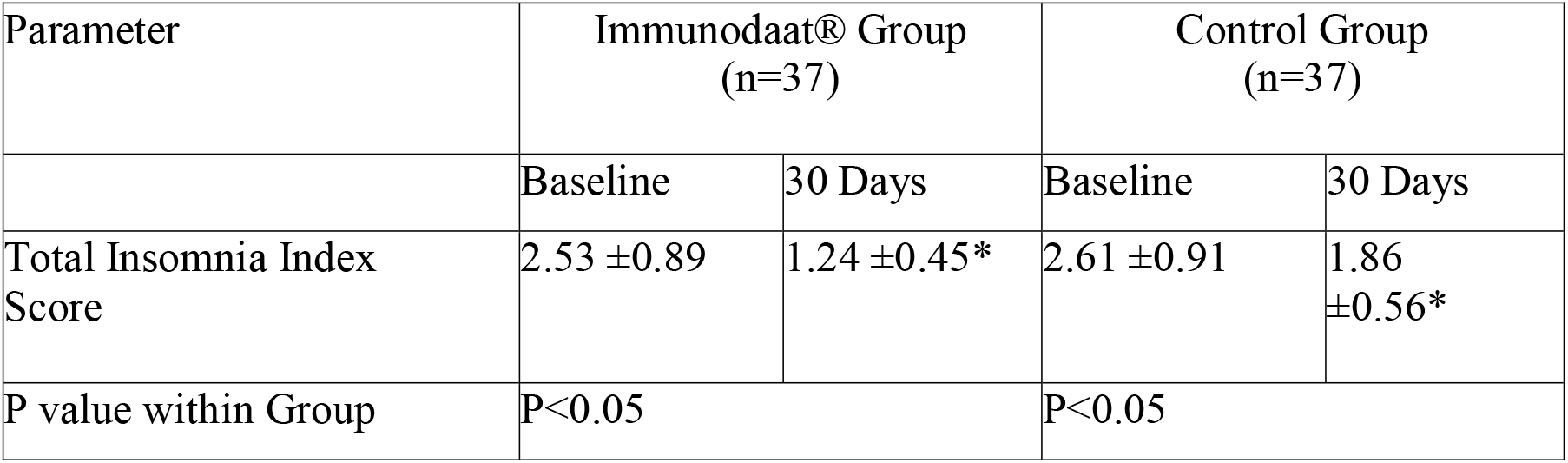
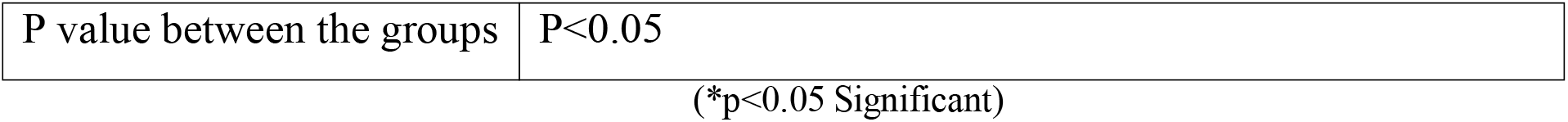
Effect on Quality of Sleep by Insomnia Severity Index (Total Score)

| Parameter | Immunodaat® Group (n=37) |  | Control Group (n=37) |  |
| --- | --- | --- | --- | --- |
|  | Baseline | 30 Days | Baseline | 30 Days |
| Total Insomnia Index Score | 2.53 ±0.89 | 1.24 ±0.45* | 2.61 ±0.91 | 1.86 ±0.56* |
| P value within Group | P<0.05 |  | P<0.05 |  |

|  |  |
| --- | --- |
| P value between the groups | P<0.05 |

Assessment of stress levels was also conducted using the perceived stress scale (PSS) on which it was observed that the levels of stress reduced from baseline to 30 days with the use ofImmunodaat® while it was not observed in the control group. Analysis between the groups also showed significantly (*p*<.05)greater stress reduction with the use of Immunodaat®. Further details in Table 6.

**Table 6:**
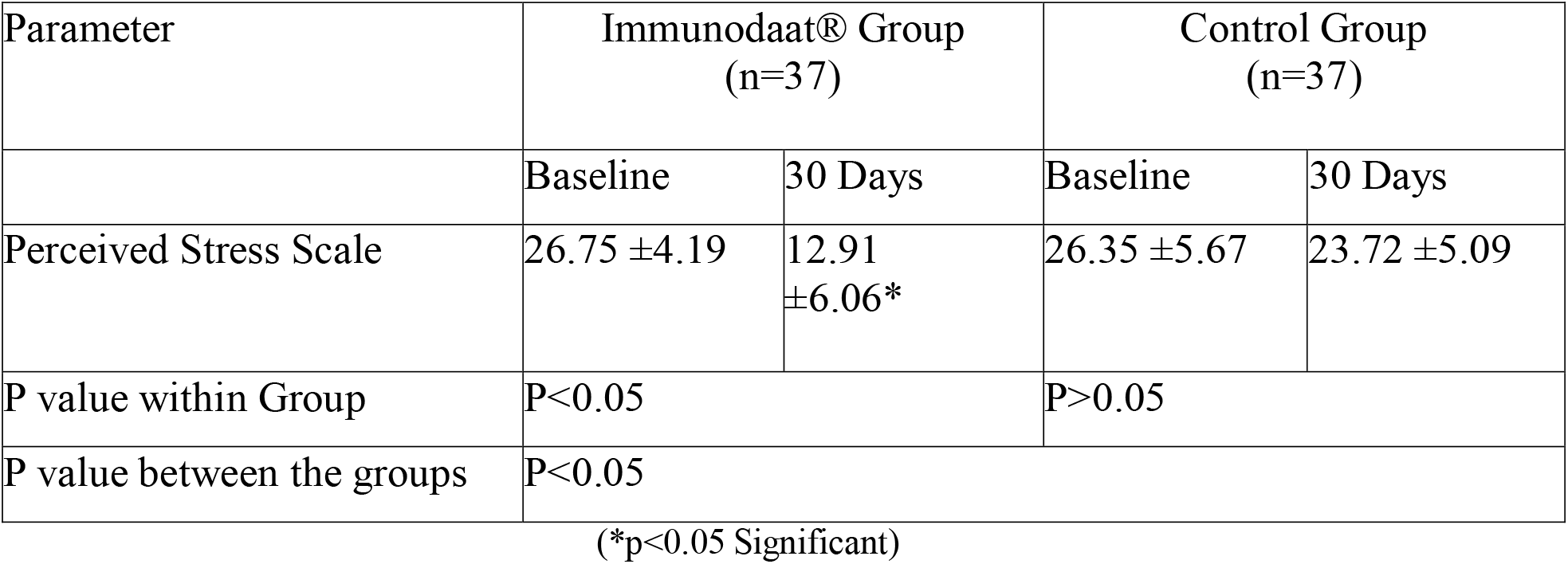
Assessment of effect on Perceived Stress Scale.

| Parameter | Immunodaat® Group<br>(n=37) |  | Control Group<br>(n=37) |  |
| --- | --- | --- | --- | --- |
|  | Baseline | 30 Days | Baseline | 30 Days |
| Perceived Stress Scale | 26.75 ±4.19 | 12.91<br>±6.06* | 26.35 ±5.67 | 23.72 ±5.09 |
| P value within Group | P<0.05 |  | P>0.05 |  |
| P value between the groups | P<0.05 |  |  |  |

(\*p<0.05 Significant)

### Assessment of effect on Quality of Life based on WHO-QOL BREF post COVID-19

Quality of life was assessed based on WHO-QOL BREF in which four primarily health related domains:physical, psychological, social, and environmental were assessed along with overall quality of life. It was observed that there was a significant improvement (*p*<.05) in the Physical & Physiological health from baseline to 30 days in Immunodaat® group as compared to control group while social health and environmental health domain did not show significant difference between the two groups. Overall quality of life score also showed a significant difference (p<0.05) from baseline to 30 days in Immunodaat® as compared to control. Refer Table 7 for details.

**Table 7:** Assessment of effect on Quality of Life based on WHO-QOL BREF.

| Parameter | Immunodaat® Group (n=37) |  | Control Group (n=37) |  | P value between groups |
| --- | --- | --- | --- | --- | --- |
|  | Baseline | 30 Days | Baseline | 30 Days |  |
| Physical Domain | 20.70± 3.95 | 25.63 ± 3.76* | 21.33 ±4.23 | 23.35 ± 3.78* | P<0.05 |
| Physiological Domain | 21.16 ± 3.86 | 24.14 ±3.11* | 21.61 ± 4.16 | 22.27 ±3.49 | P<0.05 |
| Social Domain | 8.35 ±1.97 | 9.56 ±1.55 | 10.87±1.21 | 11.60 ±1.14 | P>0.05 |
| Environmental Domain | 19.67 ±4.85 | 23.23 ±3.69* | 26.32±2.45 | 27.64 ±2.39 | P>0.05 |
| Overall Score | 3.18 ±0.60 | 4.40 ±0.67* | 3.93 ±0.66 | 4.18 ±0.72 | P<0.05 |

### Assessment of change in Energy and Stamina levels post COVID-19

Energy and stamina were assessed on the VAS scale of 0-100, where 0 indicates no energy/stamina while 100 indicates the highest level of stamina. It was observed that both energy levels and stamina levels showed significant improvement (*p*<.05) from baseline to the end of 30 days, however these parameters showed greater improvement with the use of Immunodaat® (*p*<.05). Details in Table 8.

**Table 8:** Assessment of Energy and Stamina Levels Post Covid-.

| Sr. No | Symptom | Immunodaat® Group (n=37) |  | Control Group (n=37) |  | P value between groups |
| --- | --- | --- | --- | --- | --- | --- |
|  | Parameter | Baseline | 30 Days | Baseline | 30 Days |  |
| 1 | Energy Levels | 31.35 ±6.13 | 74.57 ±8.31 | 32.13 ±8.14 | 51.87 ±5.75 | <0.05 |
|  |  | p<0.05 |  | p<0.05 |  |  |
| 2 | StaminaLevels | 33.53 ±5.48 | 80.94 ±7.43 | 35.18 ±5.80 | 58.85 ±4.15 | <0.05 |
|  |  | p<0.05 |  | p<0.05 |  |  |

### Assessment of safety

A safety assessment was performed by evaluating the incidence of adverse events and further establishing their relationship with the consumption of Immunodaat®. Assessment of laboratory parameters for complete hemogram, liver function tests, kidney function tests, lipid profile, and urine analysis wereperformed at the baseline and at the 30-day follow-up of the study.It was observed thatImmunodaat® capsules were well tolerated as they did not produce any adverse events. Also there was no significant change in the laboratory related parameters, which remained in the normal range from baseline to the end of the study. Minor adverse events were temporary andincluded abdominal discomfort and distension of abdomen. These wereresolved on their own and did not require any pause of study product or treatment. Refer to table 9 for further details.

**Table 9:** Assessment of Safety Laboratory Parameters.

| Parameter | Immunodaat® Group (n=37) |  | Control Group (n=37) |  | p-value between groups |
| --- | --- | --- | --- | --- | --- |
|  | Baseline | 30 Days | Baseline | 30 Days |  |
| COMPLETE HEMOGRAM |  |  |  |  |  |
| Hb% | 12.83 ±1.71 | 13.03 ±1.89* | 12.43±1.71 | 12.93±1.73* | p>0.05 for all values |
| RBC | 4.90 ±0.79 | 4.96 ±0.63* | 4.89 ±0.62 | 5.01 ±0.60* |  |
| WBC | 6986.48 ± 1367.31 | 6513.51 ± 1231.56* | 6713.51 ±1545.45 | 6620 ±1229.25* |  |
| Platelets | 2.51 ±0.66 | 2.34 ±0.55* | 2.32 ±0.43 | 2.52 ±0.51* |  |
| ESR | 15 ±7.23 | 13.71 ±7.78* | 11.71 ±9.55 | 12 ±7.30* |  |
| LIVER FUNCTION TESTS |  |  |  |  |  |
| Total Bilirubin | 0.53 ±0.24 | 0.60 ±0.17* | 0.65 ±0.23 | 0.61 ±0.13* | p>0.05 for all values |
| SGOT | 24.83 ±7.09 | 20.59±4.33* | 23.08±6.73 | 21.14±5.05* |  |
| SGPT | 23.24±7.66 | 22.13±4.93* | 25.89±8.72 | 24.02±5.74* |  |
| Total Proteins | 7.04 ±0.82 | 7.38 ±0.41 | 7.16 ±0.70 | 7.31 ±0.47* |  |
| AG Ratio | 1.65 ±0.43 | 1.72 ±0.41 | 1.81 ±0.54 | 1.73 ±0.29* |  |
| KIDNEY FUNCTION TESTS |  |  |  |  |  |
| Sr. Creatinine | 1.05 ±0.24 | 0.92 ±0.25 | 1.08 ±0.24 | 0.96±0.23* | p>0.05 for all values |
| BUN | 24.11 ±5.71 | 26.90±7.60 | 26.35±6.24 | 27.62±7.01* |  |

| LIPID PROFILE |  |  |  |  |  |
| --- | --- | --- | --- | --- | --- |
| Sr. Cholesterol | 189.31±16.48 | 190.10±19.94 | 187.83±23.44 | 196.20±14.85* | p>0.05<br>for all<br>values |
| Sr. Triglycerides | 118.11±34.10 | 111.43±20.63 | 115.43±29.15 | 122.67±21.40* |  |
| HDL | 38.27±8.07 | 37.10±12.23 | 38.40±7.81 | 36.18±3.89* |  |
| LDL | 126.74±13.05 | 132.75±18.21 | 128.33±22.40 | 135.48±12.11* |  |
| VLDL | 23.58±6.94 | 22.29±4.13 | 22.74±5.62 | 24.72±4.22* |  |
| TC: HDL | 5.05±0.85 | 5.44±0.48 | 5.02±0.82 | 5.46±0.48* |  |
| LDL:HDL | 3.41±0.71 | 3.80±0.48 | 3.40±0.74 | 3.78±0.48* |  |

(\*p>0.05 (Non significant difference))

## Discussion

Omicron, a mutation in SARS COV 2 is spreading faster than any previous coronavirus variant and showing signs of immune escape, with both vaccinated and previously infected people at greater risk than in previous waves. It is important to strictly follow COVID appropriate behaviors along with measures to boost immunity in order to avoid infection. Various herbal and nutraceutical supplements are available in the market to support immunity. Elderberry is one such herbal supplement that has the potential of supporting immunity andmay assist the body in supporting immunity in subjects with g influenza, and influenza-like infections. Looking at the capabilities of Elderberry, the present study was conducted to evaluate efficacy and safety of Immunodaat®Capsule that contains elderberry extract on subjects suffering from long COVID.

Thirty days of treatment with Immunodaat®resulted in a significantly reduced (*p<*.05) number of subjects with symptoms including fatigue, myalgia, weakness, joint pain, mental confusion, dizziness, alteration of mood, insomnia, and anxietywhen compared to the control group. A significant reduction (*p<*.05) was observed in the intensity of symptoms including fatigue, headache, myalgia, weakness and insomnia with the use of Immunodaat® as compared to control group. There was a significant reduction (*p<*.05)in mental health symptoms includingstress, anxiety, and difficulty concentratingin subjects treated with Immunodaat®compared to subjects in the control group. A significantly greater reduction (*p<*.05) in stress and insomnia was observed in the Immunodaat® group compared to the control group.

A significant (*p<*.05) improvement in the physical and physiological health on WHO QOL BREF was observed in the Immunodaat®group compared to the control group. Overall quality of life improved (*p<*.05) with the use of Immunodaat® as compared to the control group. It was observed that both energy levels and stamina levels showed significant improvement (*p<*.05) from baseline to the end of 30 days in both the groups, however these parameters showed greater improvement with the use of Immunodaat® capsule compared to control group.

These findings are in line with the previous discoveries on elderberry extract where in a significant reduction in severity and duration of symptoms in influenza subjects was observed.^9^A possible mechanism of action of elderberry extract in the support of immunity is the stimulation of the immune system by flavonoids enhancing the production of cytokines by monocytes.^10^ In addition, elderberry has been shown to inhibit the hemagglutination of the influenza virus and thus preventing the adhesion of the virus to the cell receptors.^11^ Anthocyanins present in elderberry also have an anti-inflammatory effect comparable to that of acetylsalicylic acid.^12-13^This could explain the pronounced effect on inflammation, pain, and fever seen in the group treated with theImmunodaat®Capsule.

It was observed thatthe Immunodaat® capsules were well tolerated by the subjects. There were minor adverse events reported, including abdominal discomfort and distension of abdomen. These were resolved on their own and did not require any pause of study product or treatment.There was no significant change in the laboratory related parameters, which remained in the normal range from baseline to the end of the study. These findings confirmed the safety of use of Immunodaat® capsules in Post COVID-19 recovery or long COVID-19.

## Conclusion

The present randomized, comparative clinical study concludes that Immunodaat®may play a significant role in supporting the body’s immune functions during symptoms of mild Post COVID-19 or long COVID. In this study, use of Immunodaat® over 30 days helped to normalize the physical and mental symptoms that occurred due to long COVID. In addition, quality of life improved with the use of Immunodaat®. These potential effects can be attributed to the active phyto-constituents having potential anti-oxidant, nutritive, nervine tonic, and adaptogenic effects ofImmunodaat®. Immunodaat®was found to be safe for consumption. Further studies should be conducted to validate these results. Key components of future studies should include a larger sample size, and blood marker evaluations..

## Data Availability

All data produced in the present work are contained in the manuscript.

## References

1. Patralekha Chatterjee. Is India missing COVID-19 deaths? The lancet. Vol 396 September 5,2020

2. Abdul Hafeez, Shmmon Ahmad, Sameera Ali Siddqui, Mumtaz Ahmad, Shruti Mishra. A Review of COVID-19 (Coronavirus Disease-2019) Diagnosis, Treatments and Prevention. EJMO 2020; 4(2):116–125

3. Clinical characteristics of COVID-19”. European Centre for Disease Prevention and Control. Retrieved 29 December 2020

4. Yelin D, Wirtheim E, Vetter P, Kalil AC, Bruchfeld J, Runold M, et al. (September 2020). “Long-term consequences of COVID-19: research needs”. The Lancet. Infectious Diseases. 20 (10): 1115–1117

5. (Filosto M, et al. J NeurolNeurosurg Psychiatry 2021;92:751–756. doi:10.1136/jnnp-2020-324837).

6. Hannah E. Davisa. Et. Al. Characterizing long COVID in an international cohort: 7 months of symptoms and their impact. Eclinicalmedicine, Volume 38, 101019, August 01, 2021

7. Trisha Greenhalgh, Matthew Knight, Christine A’Court, Maria Buxton, Laiba Husain. Management of post-acute covid-19 in primary care. BMJ 2020;370:m3026.

8. Karolina Młynarczyk, Dorota Walkowiak-Tomczak, Grzegorz P. Łysiak. Bioactive properties of Sambucus nigra L. As a functional ingredient for food and pharmaceutical industry. Journal of Functional Foods 40 (2018) 377–390

9. L. Susan Wieland, Vanessa Piechotta, Termeh Feinberg, Emilie Ludeman, Brian Hutton, Salmaan Kanji, Dugald Seely and Chantelle Garritty. Elderberry for prevention and treatment of viral respiratory illnesses: a systematic review. BMC Complementary Medicine and Therapies (2021) 21:112

10. Barak V, Halperin T, Kalickman I: The effect of Sambucol, a black elderberry-based natural product, on the production of human cytokines: I. Inflammatory cytokines. Eur Cytokine Netw 2001; 12: 290 –296.

11. Zakay-Rones Z, Varsano N, Zlotnik M, Manor O, Regev L, Schlesinger M, et al: Inhibition of several strains of influenza virus in vitro and reduction of symptoms by an elderberry extract (Sambucus nigra L.) during an outbreak of influenza B Panama. J Altern Complement Med 1995; 1: 361 –369

12. Wang H, Nair MG, Strasburg GM, Chang YC, Booren AM, Gray JI, et al: Antioxidant and antiinflammatory activities of anthocyanins and their aglycon, cyanidin, from tart cherries. J Nat Prod 1999; 62: 294 –296.

13. Christiane Schön, Yvonne Mödinger, Franziska Krüger, Cornelia Doebis, Ivo Pischel & Bernd Bonnländer (2021) A new high-quality elderberry plant extract exerts antiviral and immunomodulatory effects in vitro and ex vivo, Food and Agricultural Immunology, 32:1, 650-662,

